## Supplementary Material for "Predicting vaccine effectiveness in Mpox"

| Effectiveness Study | Vaccine | Country | Study Design | Observation period | Population/ Control | Total cases | Disaggregation[#] | Reported Doses | Included in Data Analysis^ |
|---|---|---|---|---|---|---|---|---|---|
| Breman 1980[1] | First Generation | DRC + Nigeria | Secondary Contacts | 1970-1979 | 447 | 4 | Spatial | At least 1 | Included |
| Fine 1988[2] | First Generation | DRC | Secondary Contacts | 1980-84 | 834 | 36 | Spatial | At least 1 | Not included. Major overlap with (Jezek 1988) |
| Jezek 1986[3] | First Generation | DRC | Secondary Contacts | 1980-1984 | 2510 | 56 | Spatial | At least 1 | Not included. Major overlap with (Jezek 1988) |
| Jezek 1988[4] | First Generation | DRC | Secondary Contacts | 1981-1986 | 2278 | 69 | Spatial and age | At least 1 | Included |
| Rimoin 2010[5] | First Generation | DRC | Case-Coverage | 2006-2007 | | 760 | Age | At least 1 | Included |
| Nolen 2015[6] | First Generation | DRC | Secondary contacts | 2013 | 97 | 44 | None | At least 1 | Included |
| Whitehouse 2021[7] | First Generation | DRC | Case-Coverage | 2011-2015 | | 1057 | None | At least 1 | Included |
| Wolff Sagy 2023[8] | MVA-BN | Israel | Cohort Study | 2022 | 2054 | 18 | Weekly | At least 1 | Included |
| Payne 2022a[9] | MVA-BN | US | Case-Coverage | 2022 | | 5402 | Weekly | At least 1 | Not included. Major overlap with (Payne 2022b) |
| Payne 2022b[10] | MVA-BN | US | Case-Coverage | 2022 | | 9544 | Weekly | 1 or 2 | Included |
| Bertran 2023[11] | MVA-BN | UK | Case-Coverage | 2022 | 89240 | 460 | Weekly | At least 1 | Included |

| | | | | | | | | | |
|---|---|---|---|---|---|---|---|---|---|
| Dalton* 2023[12] | MVA-BN | US | Case-Control | 2022 | 608 | 309 | None | 1 or 2 | Included |
| Deputy* 2023[13] | MVA-BN | US | Case-Control | 2022 | 8649 | 2266 | None | 1 or 2 | Included |
| Rosenberg* 2023[14] | MVA-BN | US | Case-Control | 2022 | 507 | 252 | None | 1 or 2 | Included |

Table S1: Summary of vaccine effectiveness studies identified from the systematic search and that were considered for analysis.

### Spatial disaggregation split secondary contacts between household and non-household contacts, and in some cases by proximity of household to primary case (same house, neighbouring house, other house in village or other village).

^ The indicated studies were excluded from analysis since they were highly overlapping with other studies included in the analysis and since more complete data was available from another related study.

* Dalton, Deputy and Rosenberg all use public databases of Mpox cases from similar regions and likely contain overlapping case data but due to significant differences in methodology for determining the control group, the studies were all included in the analysis.

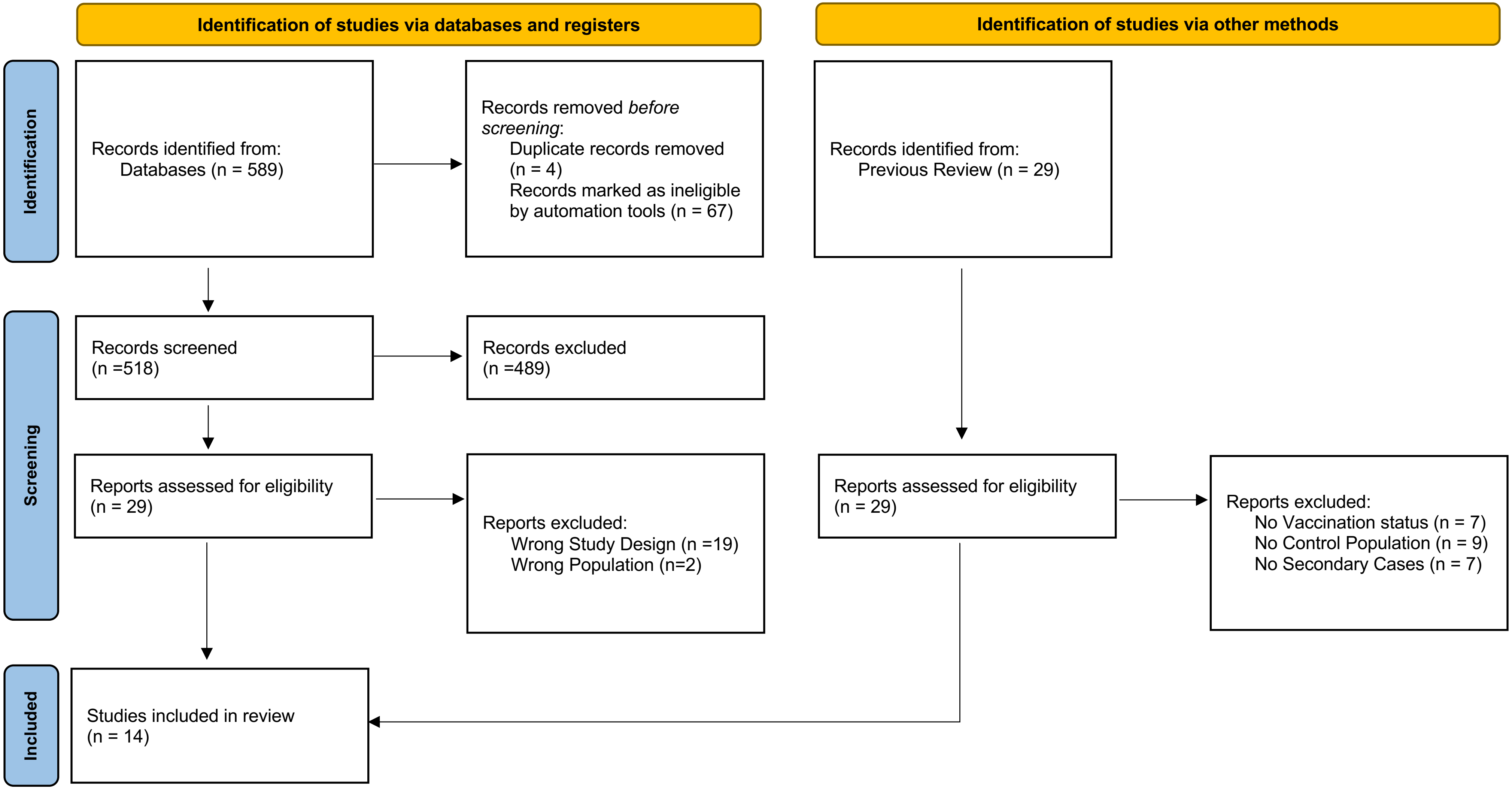


Figure S1: PRISMA flowchart for studies of the effectiveness of different Mpox vaccines identified through a systematic review process. 8 studies were included from our systematic search and 6 from a previous review.

| Clinical Trial Identifier | Participants | Prior Vaccinia Status | Medical Conditions | Doses | Age | 1 Dose MVA-BN | 2 Dose MVA-BN | ELISA Assay OD# | Formulation^ | Published article |
|---|---|---|---|---|---|---|---|---|---|---|
| NCT00189956 | 165 | Naïve | Healthy | 2 | 18-30 | Yes | Yes | 0.35 | FD | von Krempelhuber 2010[15] |
| NCT00316524 | 745 | Experienced and Naive | Healthy | 0-2 | 18-55 | Yes | Yes | NR | LF | Zitzmann-Roth 2015[16] |
| NCT01913353 | 440 | Naïve | Healthy | 2 | 18-42 | Yes | Yes | NR | LF | Pittman 2019[17] |
| NCT00316602 | 632 | Naïve | AD and healthy | 2 | 18-40 | Yes | Yes | 0.3 | LF | Greenberg 2015[18] |
| NCT00437021 | 206 | Naïve | Healthy | 1,2 | 18-38 | Yes | Yes | 0.35 | LF | Frey 2013[19] |
| NCT00316589 | 581 | Experienced and Naive | HIV and healthy | 2 | 18-55 | Yes | Yes | 0.3 | LF | Overton 2015[20] |
| NCT01144637 | 4005 | Naïve | Healthy | 2 | 18-40 | No | Yes | NR | LF | Overton 2018[21] |
| NCT00686582 | 304 | Naïve | Healthy | 3 | 18-35 | Yes | Yes | 0.35 | LF | Ilchmann 2023[22] |
| NCT00857493 | 120 | Experienced | Healthy | 2 | 56-80 | Yes | Yes | 0.3 | LF | Greenberg 2016[23] |
| NCT01668537 | 651 | Naïve | Healthy | 2 | 18-55 | Yes | Yes | NR | LF and FD | Published online only[24] |
| NCT00914732 | 523 | Naïve | Healthy | 2 | 18-38 | Yes | Yes | 0.35 | LF and FD | Frey 2015[25] |
| NCT03699124 | 1129 | Naïve | Healthy | 2 | 18-45 | No | Yes | NR | FD | Overton 2023[26] |
| NCT01827371 | 435 | Naïve | Healthy | 2 | 18-40 | Yes | Yes | 0.3 | FD | Jackson 2017[27] |

Table S2: Summary of vaccine immunogenicity trials included in our meta-analysis. Data was extracted from the clinical trials registry or, when not present in the registry, by identifying a linked research article where trial results were disseminated.

#ELISA assays were conducted at either: (a) wavelength 450nm/endpoint optical density (OD) = 0.3, or (b) wavelength 492nm/endpoint OD= 0.35. Trials where the endpoint was not reported are listed as NR.

^FD=Freeze Dried, LF=Liquid Frozen.

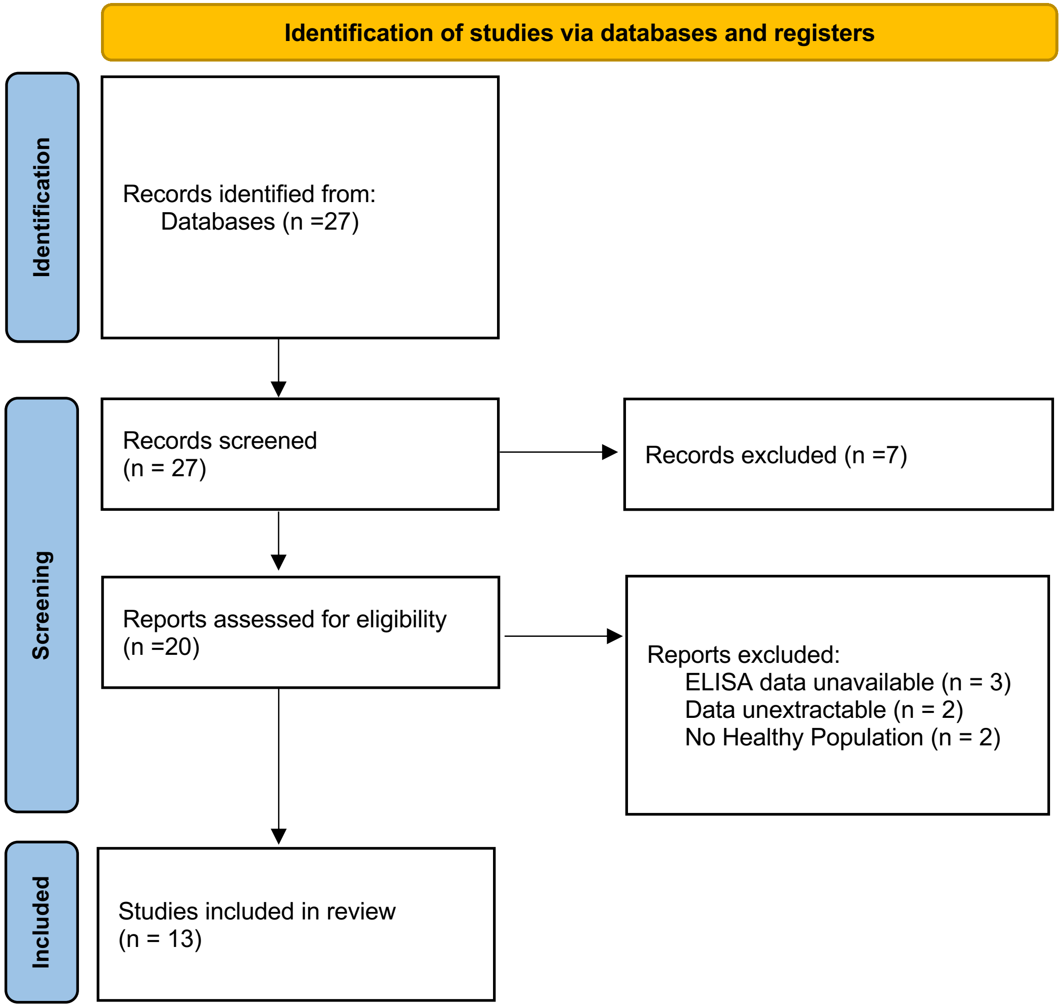


Figure S2: PRISMA flowchart for the immunogenicity studies identified as part of the systematic review process

| Parameter | | Symbol | Median (Credible Interval) | Prior |
|---|---|---|---|---|
| Effectiveness | First Generation | $E_1$ | 0.77 (0.59-0.87) | $log(1-E_1) \sim U(0,1)$ |
| | 1-Dose MVA-BN | $E_2$ | 0.74 (0.56-0.83) | $log(1-E_2) \sim U(0,1)$ |
| | 2-Dose MVA-BN | $E_3$ | 0.82 (0.69-0.89) | $log(1-E_3) \sim U(0,1)$ |
| Interstudy Standard deviation | | $\sigma_E$ | 0.46 (0.26-0.94) | Half-Cauchy(0,0.25) |

Table S3: Summary of the estimated effectiveness parameters and the prior distributions. Risk of infection $r_{i,v,s}$, and the random study effects, $S_s$, are also estimated and can be accessed using the GitHub repository.

| Parameter | | Symbol | Median (Credible Interval) | Prior |
|---|---|---|---|---|
| $\log_{10}$(GMT) | First Gen | $\mu_1$ | 1.77 (1.61-1.91) | Normal(0,10) |
| | 1-Dose MVA-BN | $\mu_2$ | 1.94 (1.82-2.06) | Normal(0,10) |
| | 2-Dose MVA-BN | $\mu_3$ | 2.87 (2.75-2.98) | Normal(0,10) |
| Standard deviation of ($\log_{10}$) titers | First Gen | $\sigma_1$ | 0.765 (0.710-0.827) | Log-normal(0,10) |
| | 1-Dose MVA-BN | $\sigma_2$ | 0.662 (0.643-0.683) | Log-normal(0,10) |
| | 2-Dose MVA-BN | $\sigma_3$ | 0.386 (0.378-0.394) | Log-normal(0,10) |
| Formulation Effect | | $\mu_f$ | 0.13 (0.08-0.17) | Normal(0,1) |
| ELISA effect | | $\mu_E$ | 0.14 (-0.27-0.54) | Normal(0,1) |
| Interstudy Standard Deviation | | $\sigma_I$ | 0.17 (0.11-0.30) | Half-Cauchy(0,1) |

Table S4: Summary of the estimated parameters from fitting the immunogenicity data. The study effects are not included in the table.

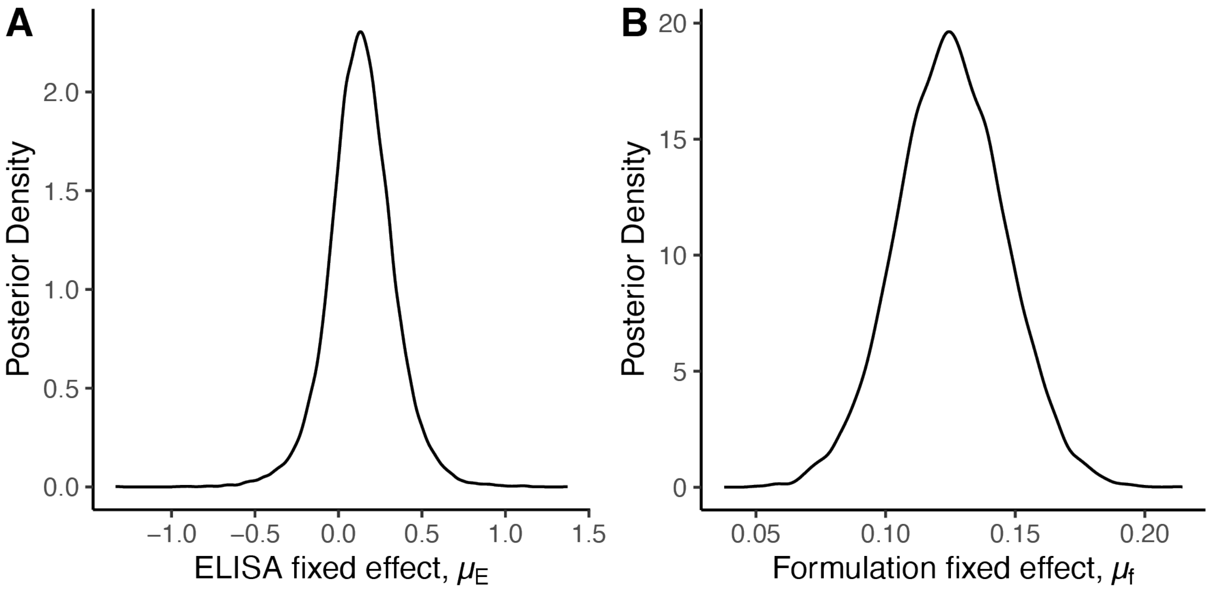


Figure S3: The posterior distributions of the fixed effects on the ($\log_{10}$) GMT in the immunogenicity model. (A) The effect of the ELISA assay was not found to be significant (fold-difference ($10^{\mu_E}$) 1.3-fold, CI:0.54-3.48). (B) The effect due to different formulations of MVA-BN was found to be significant (fold-difference ($10^{\mu_f}$) 1.33-fold CI:1.21-1.46).

| Parameter | | Symbol | Estimated Value |
|---|---|---|---|
| $\log_{10}$(GMT) | First Gen | $\mu_1$ | 1.77 (1.62-1.91) |
| | 1-Dose MVA-BN | $\mu_2$ | 1.94 (1.83-2.05) |
| | 2-Dose MVA-BN | $\mu_3$ | 2.87 (2.76-2.98) |
| Standard deviation of ($\log_{10}$) titers | First Gen | $\sigma_1$ | 0.77 (0.72-0.84) |
| | 1-Dose MVA-BN | $\sigma_2$ | 0.67 (0.65-0.69) |
| | 2-Dose MVA-BN | $\sigma_3$ | 0.39 (0.38-0.40) |
| Formulation Effect | | $\mu_f$ | 0.12 (0.08-0.17) |
| Logistic Slope | | $k$ | 0.47 (0.19-0.76) |
| Logistic Constant | | $A$ | 1.49 (1.03-1.96) |
| Interstudy Standard Deviation | Immunogenicity | $\sigma_I$ | 0.17 (0.11-0.30) |
| | Efficacy | $\sigma_E$ | 0.42 (0.25-0.82) |

Table S5: The estimated parameters from the fitted logistic model relating data on vaccine immunogenicity and effectiveness (Figure 3).

| Parameter | | Symbol | Prior |
|---|---|---|---|
| $\log_{10}$(GMT) | First Gen | $\mu_1$ | $\mathrm{Normal}(0,10)$ |
| | 1-Dose MVA-BN | $\mu_2$ | $\mathrm{Normal}(0,10)$ |
| | 2-Dose MVA-BN | $\mu_3$ | $\mathrm{Normal}(0,10)$ |
| Standard deviation of ($\log_{10}$) titers | First Gen | $\sigma_1$ | $\mathrm{Log\text{-}normal}(0,10)$ |
| | 1-Dose MVA-BN | $\sigma_2$ | $\mathrm{Log\text{-}normal}(0,10)$ |
| | 2-Dose MVA-BN | $\sigma_3$ | $\mathrm{Log\text{-}normal}(0,10)$ |
| Formulation Effect | | $\mu_f$ | $\mathrm{Normal}(0,1)$ |
| Logistic Slope | | $k$ | $\mathrm{Normal}(0,1)$ |
| Logistic Constant | | $A$ | $\mathrm{Logistic}(0,1)$ |
| Interstudy Standard Deviation | Immunogenicity | $\sigma_I$ | $\mathrm{Half\text{-}Cauchy}(0,1)$ |
| | Efficacy | $\sigma_E$ | $\mathrm{Half\text{-}Cauchy}(0,0.25)$ |
| Risk of Vaccination/Infection | | $r_{i,v,s}$ | $\mathrm{Beta}(1,1)$ |
| Study Effects | Immunogenicity | $\mu_I$ | $\mathrm{Normal}(0,\sigma_I)$ |
| | Efficacy | $\mu_E$ | $\mathrm{Normal}(0,\sigma_E)$ |

Table S6: List of all the model parameters and priors used to fit a logistic relationship between efficacy and immunogenicity data (Figure 3). The study effects are model parameters within a hierarchical structure and the reported prior is the modelled hierarchy.

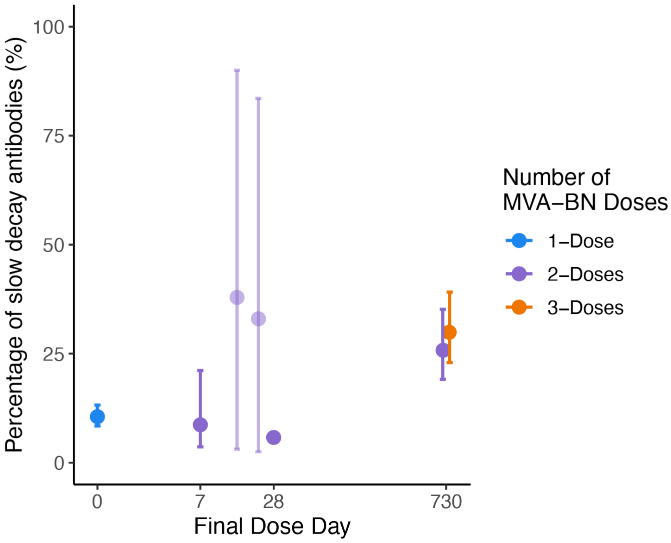


Figure S4: The proportion of slow-decaying (long-lived) antibodies induced from vaccination with different dosing schedules. Primary doses are administered on day 0, with the three-dose schedule including a second dose 28-days after the primary dose. Percentages for regimens without a datapoint later than 5 months post-vaccination are faded.

| Model | LPPD | $P_{waic}$ | WAIC |
|---|---|---|---|
| Two Phase Decay Model with distinct decay rates | -1756.6 | 199.6 | 3912 |
| Two Phase Decay Model with common decay rates | -1756.9 | 196.8 | 3907 |
| Single Phase Decay Model with distinct decay rates | -2435.4 | 744.0 | 6359 |
| Single Phase Decay Model with common decay rate | -2676.1 | 683.3 | 6718 |
| No Decay Model | -3422.1 | 839.1 | 8522 |

Table S7: Comparison of the Widely Applicable Information Criterion (WAIC) for different models of the decay in antibody binding. Higher log-posterior predictive density (LPPD) indicates a greater predictive fit. The $P_{waic}$ is a penalty term.

| Vaccination Schedule | Predicted ELISA GMT (95% Credible Interval) at time points: | | | | |
|---|---|---|---|---|---|
| | 3 months | 6 months | 12 months | 18 months | 24 months |
| 1-Dose | 12 (9-15) | 8 (6-11) | 7 (5-10) | 7 (5-9) | 6 (5-9) |
| 2-Dose (Day-28) | 68 (52-91) | 38 (30-48) | 34 (26-43) | 31 (24-41) | 29 (22-39) |
| 2-Dose (Day-730) | 610 (451-834) | 517 (375-720) | 476 (342-670) | 441 (314-633) | 410 (286-601) |
| 3-Dose | 714 (535-959) | 619 (458-844) | 571 (419-785) | 530 (384-742) | 493 (347-705) |

Table S8: The predicted vaccinia-binding GMT at different time points after the peak titer. We compare the effect of different vaccination schedules on the expected titer.

**Supplementary Methods:**

**Derivation of Binomial model for case-control studies:**

Let $P(V|I) = \phi$ be the probability that a person is vaccinated (V) given they are infected (I). Then,

$$P(I|V) = \frac{P(V|I)P(I)}{P(V)}.$$

The calculation for $P(I|U)$ (Probability of infection given unvaccinated (U)), is similar and we can then calculate the ratio,

$$\frac{P(I|V)}{P(I|U)} = \frac{P(V|I)}{P(U|I)}\frac{P(U)}{P(V)}.$$

Conducting a similar calculation for the control group, let $P(V|C) = \gamma$, where $C$ is the control group, then,

$$1 - E = \frac{P(I|V)}{P(I|U)} = \frac{\phi}{1-\phi}\frac{1-\gamma}{\gamma}\frac{P(C|V)}{P(C|U)}. \qquad \text{S1}$$

In an ideal case-control study design the fraction $\frac{P(C|V)}{P(C|U)} = 1$. However, unmeasured confounding can cause this quantity to vary around 1. Since this cannot be quantified directly, we incorporate this ratio into a random effect with $S = \ log\frac{P(C|U)}{P(C|V)}$. Taking logarithms of Equation S1 we obtain,

$$logit(\phi) = logit(\gamma) + \log\,(1-E) + S.$$

This is equivalent to a model where the relative risk reduction of the vaccine has a random effect between studies. Comparing this model to the model used for case-coverage studies (equation 1 of the Methods) we find that both study types are described by applying the same model, except here $r_{v,0,s} = \gamma$, and $r_{v,i,s} = \phi$, whereas in the case-coverage method (from equation 1 of Methods), $r_{i,0,s} = \gamma$, and $r_{i,v,s} = \phi$ . That is, for case-control studies,

$$\text{logit}(r_{v,i,s}) = \text{logit}(r_{v,0,s}) + \log(1-E_v) + S_s, \qquad S2$$

where $r_{v,0,s}$ and $r_{v,i,s}$ denote the probability of being vaccinated in the control and infected cohorts, respectively. The indexation in this case differs slightly from the case-coverage methods. That is, here the different disaggregation groups correspond to the vaccine ($v$), while the control and infectious groups are indexed by ($i$).

**Immunogenicity data:**

The extracted immunogenicity data included geometric mean titers and confidence intervals. Using the extracted confidence intervals, we calculate the approximate sample standard deviation, $s$, from each study and group assuming confidence intervals were calculated using the formula,

$$\mathrm{CI} = \log_{10}(GMT) \pm t_{0.05,n}\frac{s}{\sqrt{n}},$$

where n is the sample size, GMT is the reported geometric mean titer, $t_{0.025,n}$ is the 0.025 percentile of the t-distribution with n-degrees of freedom. Due to rounding of the reported values, using the upper or lower confidence limits yielded different results. Hence, we average across the sample standard deviation estimates calculated from the upper and lower confidence limits.

In our evaluation of the posterior distribution, we used the sample mean ($\log_{10}$) antibody titer, $\bar{y}_{s,v,f,E}$, and sample standard deviation of these ($\log_{10}$ titers), $\overline{sd}_{s,v,f,E}$ (for each vaccine, $v$, from each study, $s$, with vaccine formulation, $f$, and ELISA assay, $E$), rather than the individual titers (individual titers were not provided upon request). These are sufficient statistics (as will be seen below) to define our distribution of interest.

For simplicity, let $\mu_{s,v,f,E}$ denote the mean of a cohort from a study $s$, using the vaccine $v$, the ELISA $E$, and formulation $f$. That is, $\mu_{s,v,f,E} = \mu_v + \mu_E + \mu_f + \mu_s$, where, $\mu_v$, is the mean of the 'true' distribution of titers (administered with liquid frozen formulation), $\mu_E$, is the effect of the ELISA assay on the observed titers, $\mu_f$, is the effect of using a freeze dried formulation of the vaccine and $\mu_S$, is the random study effect on the observed titers. Then the likelihood of observing a particular set of individuals ($\log_{10}$) antibody titers, $\boldsymbol{y}_{s,v,f,E}$, from a particular study/group/etc, given a certain (as yet to be estimated) $\mu_{s,v,f,E}$ and $\sigma_v$ (standard deviation of the titers from the vaccine group) is given by,

$$p\left(\boldsymbol{y}_{s,v,f,E} \middle| \mu_{s,v,f,E}, \sigma_{\mathrm{v}}\right) \propto \prod_i \frac{1}{\sigma_v\sqrt{2\pi}} \exp\left(\frac{-1}{2}\left(\frac{\mathrm{y_i} - \mu_{s,v,f,E}}{\sigma_v}\right)^2\right)$$

where $y_i$ are the individual titers of $\boldsymbol{y}_{s,v,f,E}$. Rearranging this equation, we can write this as,

$$p\left(\boldsymbol{y}_{s,v,f,E}\middle|\mu_{s,v,f,E},\sigma_v\right) \propto \frac{1}{\sigma_v^n}\exp\left(\frac{-1}{2\sigma_v^2}\sum_i\left(y_i-\mu_{s,v,f,E}\right)^2\right).$$

Constant terms can be ignored as we only require a proportional function to sample from the posterior[28]. Focusing on the summation in the exponent and using the definition of a sample standard deviation and sample mean, we can expand the summation and simplify obtaining,

$$\sum_i\left(y_i-\mu_{s,v,f,E}\right)^2 = \left(n_{s,v,f,E}-1\right)\overline{sd}^2 + n_{s,v,f,E}\left(\bar{y}_{s,v,f,E}-\mu_{s,v,f,E}\right)^2,$$

where $n_{s,v,f,E}$ is the number of individual titers used in the sample mean and standard deviation for the given vaccine, study, formulation and ELISA combination. It follows that we only need the sample mean, $\bar{y}_{s,v,f,E}$, and sample standard deviation, $\overline{sd}_{s,v,f,E}$, as sufficient statistics, to evaluate the likelihood function. The contributions of each cohort are multiplied together, yielding the full likelihood,

$$p\left(\bar{y}_{s,v,f,E},\overline{sd}_{s,v,f,E}\middle|\mu_{s,v,f,E},\sigma_v,n_{s,v,f,E}\right)$$
$$\propto \prod \frac{1}{\sigma_v^{n_{s,v,f,E}}}exp\left(\frac{-1}{2\sigma_v}\left(\left(n_{s,v,f,E}-1\right)\overline{sd}_{s,v,f,E}+n_{s,v,f,E}\left(\bar{y}_{s,v,f,E}-\mu_{s,v,f,E}\right)\right)\right)$$

The posterior distribution for all the parameters can then be determined using this likelihood function in conjunction with the priors (specified in the main text Methods).
